# Comparison of two automated CT perfusion software packages in patients with ischemic stroke presenting within 24 hours of onset

**DOI:** 10.1101/2024.02.12.24302732

**Authors:** Nak-Hoon Kim, Sue Young Ha, Gihoon Park, Jong-Hyeok Park, Dongmin Kim, Leonard Sunwoo, Min-Surk Kye, Sung Hyun Baik, Cheolkyu Jung, Wi-Sun Ryu, Beom Joon Kim

## Abstract

**Background:** We compared the ischemic core and hypoperfused tissue volumes estimated by RAPID and JBS-10K, a newly developed automated CT perfusion (CTP) analysis package. We also assessed agreement between ischemic core volumes by two software packages against early follow-up infarct volumes on diffusion-weighted images (DWI).

**Methods:** This retrospective study analyzed 327 patients admitted to a single stroke center in Korea from January 2021 to May 2023, who underwent CTP scans within 24 hours of onset. The concordance correlation coefficient and Bland-Altman plots were utilized to compare the volumes of ischemic core and hypoperfused tissue volumes between the software packages. Agreement with early (within 3 hours from CTP) follow-up infarct volumes on diffusion-weighted imaging (n=217) was also evaluated.

**Results:** The mean age was 70.7±13.0 and 137 (41.9%) were female. Ischemic core volumes by JBS-10K and RAPID in the threshold of relative cerebral blood flow (rCBF) < 30% had excellent agreement (ρ = 0.958 [95% CI, 0.949 to 0.966]). Excellent agreement was also observed for time to a maximum of the residue function (Tmax) > 6 seconds between JBS-10K and RAPID (ρ = 0.835 [95% CI, 0.806 to 0.863]). Although early follow-up infarct volume showed substantial agreement in both packages (JBS-10K ρ = 0.751 and RAPID ρ = 0.632), ischemic core volumes at the threshold of rCBF <30% tended to overestimate ischemic core volumes.

**Conclusion:** JBS-10K and RAPID demonstrated remarkable concordance in estimating the volumes of the ischemic core and hypoperfused area based on CTP within 24 hours from onset.

## INTRODUCTION

In the rapidly evolving field of neuroimaging, the analysis of computed tomography perfusion (CTP) scans has become a cornerstone in the diagnosis and management of acute ischemic stroke.^1^ CTP scans have played a major role in expanding the time window for endovascular treatment (EVT) in patients with ischemic stroke.^2,3^ Using perfusion scans, clinical trials that compare EVT with medical treatment for patients with anterior circulation large vessel occlusion beyond 6 hours have shown the clinical benefit of EVT in the extended time window.^2,3^ Furthermore, our group has demonstrated that even beyond 24 hours, selected individuals identified through perfusion imaging can benefit from EVT.^4^ In this context, It is of the utmost importance to precisely quantify the perfusion parameters to make an informed decision for acute ischemic stroke patients.^5^

To streamline the analysis of perfusion and minimize variations among different observers, various commercial CTP software solutions have been introduced.^6^ These solutions automatically identify the ischemic core and penumbral regions. Nonetheless, there has been considerable inconsistency in the parameters of CTP and the quantitative benchmarks set for delineating the ischemic core and penumbra.^7^ Gradually, relative CBF (rCBF) has emerged as the preferred metric for determining the ischemic core.^8^ A threshold for the time to peak of the residue function (Tmax) exceeding 6 seconds has been identified as a reliable predictor for tissues at risk of infarction if there is no reperfusion.^9^ However, the extent to which different software solutions can be used interchangeably, especially in terms of their clinical significance for planning treatment and estimating prognosis, remains uncertain. Previous research has indicated significant discrepancies in the calculated volumes of the ischemic core across different software, leading to inconsistent predictions of final infarct volume after mechanical thrombectomy.^7,10^

Previous research has shown that the epidemiology of stroke, encompassing its frequency, contributing factors, and death rates, differs markedly across ethnic groups.^11–13^ For example, in comparison to North Americans and Europeans, the incidence of intracranial arterial disease is comparatively higher among Asians and Blacks.^12^ One may postulate that ischemic core thresholds derived from studies on Caucasian populations may lead to an overestimation of ischemic core volumes in Asian patients due to the pre-stroke development of leptomeningeal collaterals after chronic intracranial stenosis.^14,15^ Additionally, disparities in ischemic core volume may be influenced by various comorbidities, such as hypertension and diabetes, as well as delays in both seeking and receiving timely stroke treatment.

In this study, we compared the volumes of ischemic core and hypoperfused tissue volumes estimated by RAPID (iSchemaView Inc, Menlo Park, CA) and JBS-10K (JLK Inc., Seoul, Korea). In addition, we aimed to evaluate the agreement of patient selection eligible for EVT between the two packages.

## METHODS

### Study design and Study population

In this retrospective study using prospectively collected data, we included patients who were admitted to Seoul National University Bundang Hospital between January 2021 and May 2023 and underwent CTP scans within 24 hours of symptom onset. We excluded patients with 1) severe motion artifacts on CTP or poor contrast bolus arrival and 2) failed automated perfusion calculation by RAPID. A subset of patients with available follow-up diffusion-weighted image (DWI) underwent within 3 hours from CTP and was used to compare the baseline ischemic core volumes predicted by the CTP with the early follow-up infarct volumes.

The study protocol was approved by the institutional review board of Seoul National University Bundang hospital [IRB# B-1710-429-102], and patients or their legal representatives provided a written informed consent.

### Clinical data collection

Using a standardized protocol,^16^ we prospectively collected demographic data, prior medication history, and the presence of vascular risk factors including hypertension, diabetes mellitus, hyperlipidemia, coronary artery disease, atrial fibrillation, and smoking history. Stroke subtypes were determined by an experienced vascular neurologist, using a validated MRI-based classification system built on the TOAST criteria.^17^

### Imaging and image reconstruction

All CTP scans were performed using a 256-Slice CT scanner (Brilliance iCT 256, Philips Healthcare, Best, the Netherlands). The imaging parameters for CTP scans were as follows: 80 kVp, 150 mAs, Beam collimation 6×1.25 mm, rotation time 0.45 seconds. In the arterial phase scan, 30 cycles were captured every 2 seconds, resulting in a total duration of 60 seconds. Following a delay of 9 seconds, the delayed phase scan acquires 2 cycles at intervals of 10 seconds, amounting to a total duration of 20 seconds. A total of 50 mL of iodinated contrast agent (Iomeprol 400; Bracco Imaging, Milan, Italy) followed by 30 mL of saline flush was administered intravenously at a rate of 5 mL/s. The images were reconstructed with the slice thickness of 10 mm and the increment of 10 mm covering 8 cm in z-axis

### Automated software analyzing CTP

JBS-10K is a fully automated CTP software package to visualize and quantify ischemic core and hypoperfused tissue, which relies on a block-circulant singular value decomposition (cSVD) method.^18^ We compared the volumes of ischemic core and hypoperfused tissue volumes estimated by RAPID and JBS-10K, both of which are automated CTP analysis packages using a delay-insensitive algorithm. A default setting of rCBF < 30% was used for defining the ischemic core. For assessing of hypoperfused tissue volume, we used the default setting of both packages (Tmax > 6s). Both software packages carry out automated registration, segmentation, and motion correction, employing a delay-insensitive method, and conducting post-processing (Figure 1). The RAPID software package has been routinely used for CTP in our hospital setting. Restored CTP source images were analyzed with the research version of JBS-10K within the hospital for this study.

**Figure 1.**
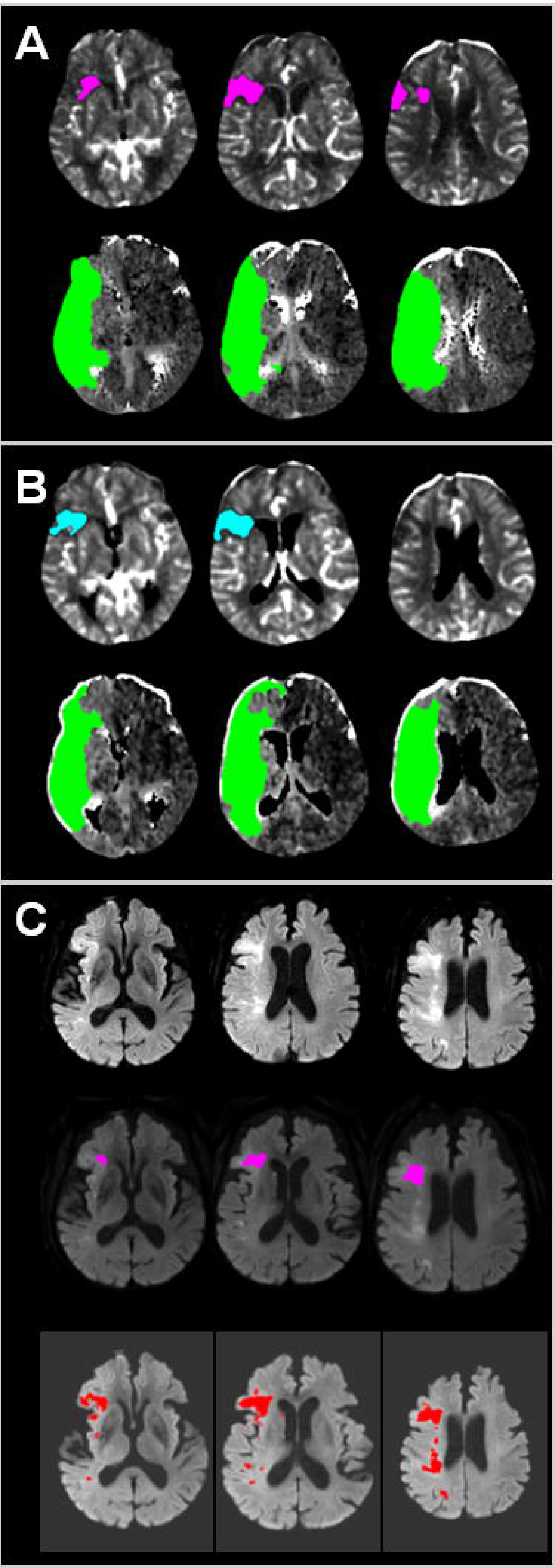
A representative case employing JBS-10K and RAPID. (A) Color map produced by RAPID, indicating an ischemic core volume (rCBF < 30%) of 10 ml and total hypoperfused tissue (Tmax > 6 seconds) of 168 ml. (B) Color map generated by JBS-10K, showing an ischemic core volume (rCBF < 30%) of 8.8 ml and total hypoperfused tissue (Tmax > 6 seconds) of 159.1 ml. (C) Early follow-up DWI taken 110 minutes after CTP, following endovascular treatment with TICI 2a, reveals patchy areas of abnormally restricted diffusion in territories of the right middle cerebral artery (Top). RAPID DWI, based on the threshold of the apparent diffusion coefficient, estimated a total infarct volume of 3 mL (Middle). JBS-01K, utilizing a deep learning algorithm, estimated a total infarct volume of 14.61 mL (Bottom). CTP = computed tomography perfusion; rCBF = relative cerebral blood flow; Tmax = time to maximum of the residue function; DWI = diffusion-weighted image.

### Early follow-up infarct volume measurement

We collected DWI taken within 3 hours of the CTP scans. Areas of infarction were automatically segmented using a validated deep learning algorithm (JBS-01K, JLK Inc., Seoul, Korea).^19^ Automatically segmented infarct areas underwent visual inspection by a stroke neurologist (W-S. Ryu) and were manually corrected as necessary.

### Statistical analysis

Data were presented as mean ± standard deviation, median (interquartile range), and number (percentage), as appropriate. To compare the volumes of ischemic core and hypoperfused tissue using two software packages, we utilized the concordance correlation coefficient (ρ) with 95% confidence intervals (CIs) and further investigated the data using reduced major axis regression.^20^ The magnitude of agreement was classified as follows: values from 0.0 to 0.2 indicating poor agreement; 0.21 to 0.40 indicating fair agreement; 0.41 to 0.60 indicating moderate agreement; 0.61 to 0.80 indicating substantial agreement; and 0.81 to 1.0 indicating excellent agreement. Additionally, Bland-Altman plots were used to assess the agreement of ischemic core volumes and hypoperfused areas as determined by the two packages. For the comparison between follow-up infarct volumes and ischemic core volumes analyzed by the two packages, both concordance correlation coefficients and Bland-Altman plots were employed. In the subgroup analysis, concordance correlation coefficients and Bland-Altman plots were utilized after stratifying patients by endovascular treatment and early infarct volume, arbitrarily divided at 20 mL. The agreement between ischemic core volume and follow-up infarct volume, categorized according to the DAWN (DWI or CTP Assessment with Clinical Mismatch in the Triage of Wake-Up and Late Presenting Strokes Undergoing Neurointervention with Trevo) clinical trial’s criteria,^2^ was assessed using Cohen’s kappa. A two-tailed *P* value of < 0.05 was considered statistically significant. All statistical analyses were performed using STATA (version 16.0, StataCorp LP, College Station, TX).

## RESULTS

### Baseline characteristics of the study population

During the study period, a total of 2,544 patients were diagnosed with ischemic stroke, and 657 patients underwent a CTP scan. We excluded 315 patients who received CTP more than 24 hours after their last known well and 15 patients who had severe motion artifacts or whose automated perfusion calculations failed using RAPID. Among the 327 patients included in the analysis, the mean age was 70.7 (SD 13.0), and 41.9% were women (Table 1). The median of the initial NIH stroke scale score was 9 (interquartile range [IQR] 4 to 17). The most common stroke etiology was identified as cardioembolism. Of the 205 (62.7%) patients who received revascularization therapy, 83 (25.3%) underwent intravenous thrombolysis alone, 59 (18.0%) underwent endovascular treatment alone, and 63 (19.3%) received combined therapy. The median interval between the time last known well to CTP scan was 192 min (IQR, 101 to 395 min). The median interval between CTP and DWI was 41 min (IQR, 27 to 100 min).

**Table 1.** Baseline characteristics.

|  |  |
| --- | --- |
| Variables | (N = 327) |
| Age, year | 70.7±13.0 |
| Sex, women | 137 (41.9%) |
| Initial NIHSS score | 9 (4 - 17) |
| Pre-stroke mRS 2 or less | 279 (88.4%) |
| Previous stroke history | 71 (21.7%) |
| Hypertension | 229 (70.0%) |
| Diabetes | 117 (35.8%) |
| Hyperlipidemia | 163 (49.9%) |
| Smoking | 79 (24.2%) |
| Atrial fibrillation | 110 (33.6%) |
| Stroke subtype |  |
| Large artery atherosclerosis | 104 (34.0%) |
| Small vessel occlusion | 27 (8.8%) |
| Cardioembolism | 112 (36.6%) |
| Undetermined | 41 (12.5%) |
| Other determined | 22 (7.2%) |
| Revascularization therapy |  |
| Intravenous thrombolysis | 83 (25.4%) |
| Endovascular treatment | 59 (18.0%) |
| Combined | 63 (19.3%) |
| Time data |  |
| Last known well to arrival, min | 147 (73 - 292) |
| Arrival to CTP, min | 28 (24 - 38) |
| CTP to DWI, min (n = 218) | 41 (27 - 100) |

Data were presented as mean ± SD, number (percentage), or median (interquartile range).
NIHSS=National Institute of Health Stroke Scale; mRS=modified Rankin Scale;
CTP=compute tomography perfusion; DWI=diffusion-weighted image.

### Concordance correlation analysis of the volumes of ischemic core and hypoperfused tissue by RAPID vs. JBS-10K

The mean difference between ischemic core volumes calculated by RAPID and JBS-10K was −0.54 mL (95% CI, −1.72 to 0.64 mL; P = 0.85). There was excellent agreement in between JBS-10K and RAPID (ρ = 0.958 [95% CI, 0.949 to 0.966]; Figure 2A). In the Bland-Altman plot, the limits of agreement were −20.74 and 21.83 mL (Figure 2B). We observed excellent agreement between JBS-10K and RAPID (ρ = 0.942 [95% CI, 0.916 to 0.960]) with the mean difference of −0.94 mL (95% CI, - 4.90 to 3.00 mL), when we restricted the analysis to patients whose ischemic core volume was not zero by both JBS-10K and RAPID (n = 90; Supplementary Figure 1).

**Figure 2.**
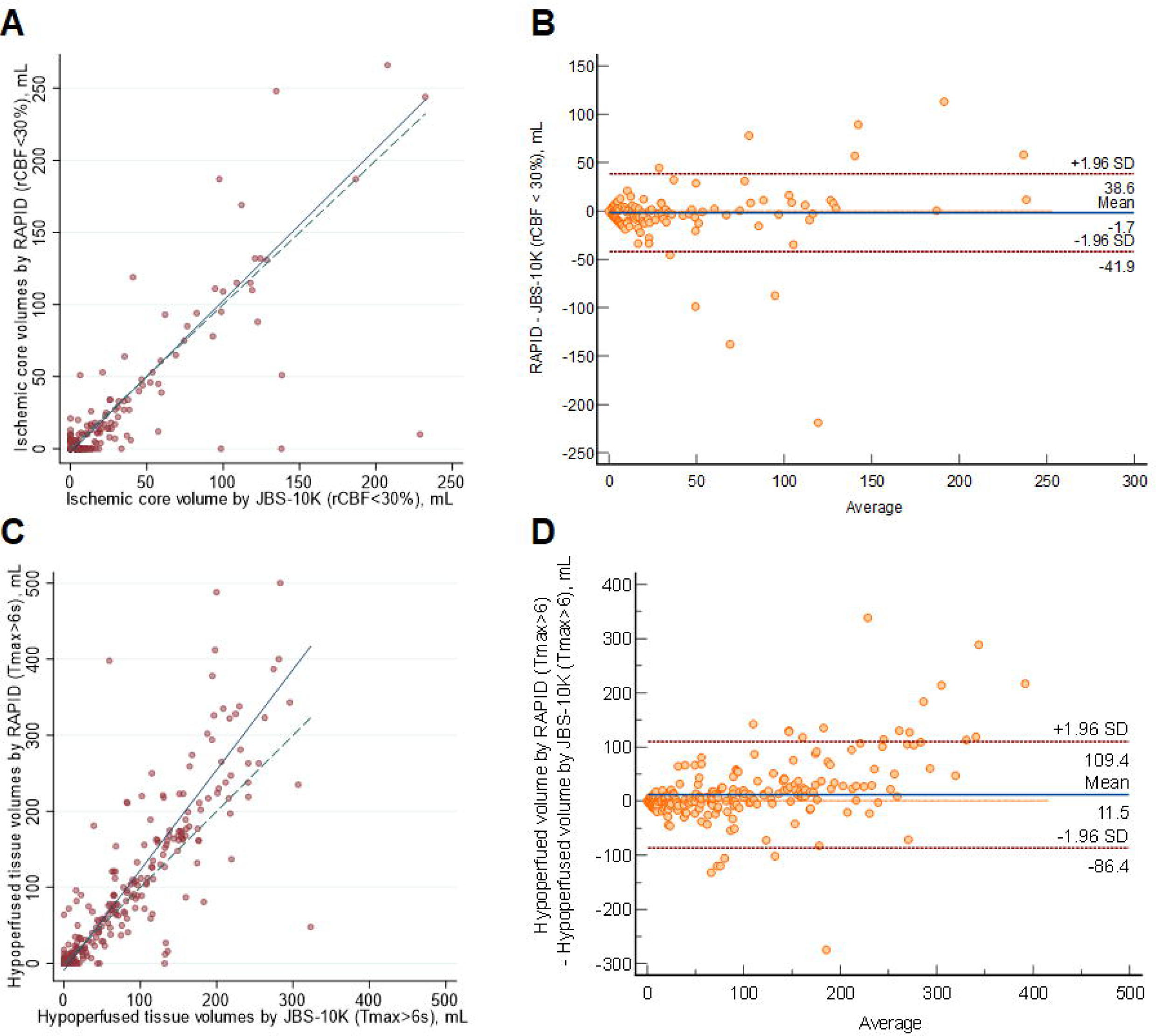
Comparison of ischemic core volumes and hypoperfused tissue volumes by JBS-10K and RAPID. (A) Scatter plot illustrating ischemic core volumes as determined by JBS-10K and RAPID, with the slope and intercept of the reduced major axis being 1.097 and −2.067, respectively. (B) Bland-Altman plot for the analysis of agreement in ischemic core volumes. (C) Scatter plot illustrating hypoperfused tissue volumes as determined by JBS-10K and RAPID, with the slope and intercept of the reduced major axis being 1.337 and −8.350, respectively. (D) Bland-Altman plot for the analysis of agreement in hypoperfused tissue volumes. The green dotted line represents the line of perfect concordance, while the blue line denotes the reduced major axis. For B and D, the blue solid line and the red dotted lines represent the mean difference and the limits of agreement between JBS-10K and RAPID, respectively.

The mean difference between hypoperfused tissue volumes calculated by JBS-10K and RAPID was 13.31 mL (95% CI, 8.22 to 18.39; P = 0.06). There was excellent agreement between JBS-10K and RAPID (ρ = 0.855 [95% CI, 0.828 to 0.877]; Figure 2C). In the Bland-Altman plot, the limits of agreement were −78.22 and 104.83 mL (Figure 2D). When the analysis was confined to patients receiving EVT (n = 122), there was excellent agreement in ischemic core volumes and substantial agreement in hypoperfused tissue volumes between JBS-10K and RAPID (Supplementary Figure 2).

### Comparison of ischemic core volumes calculated by software tools and follow-up infarct volumes on diffusion-weighted images

In patients with available early follow-up DWI (n = 218), there was substantial agreement between follow-up infarct volumes and ischemic core volumes as determined by JBS-10K (ρ = 0.753 [95% CI, 0.699 to 0.800]; Figure 3A) and RAPID (ρ = 0.736 [95% CI, 0.685 to 0.780]; Figure 3C) at the default rCBF threshold of <30%. The limits of agreement for the volumes of the ischemic core and the follow-up infarct volume were comparable between the two packages (Figures 3B and 3D). Nevertheless, at the default rCBF threshold of <30%, both packages tended to overestimate infarct core volumes, as indicated by the slope of the reduced major axis being under one. Furthermore, in patients with visible infarcts on the follow-up DWI (n = 187), the ischemic core volume determined by RAPID was zero in 123 (65.8%) cases, whereas it was zero in 104 (55.6%; P = 0.04) cases as determined by JBS-10K.

**Figure 3.**
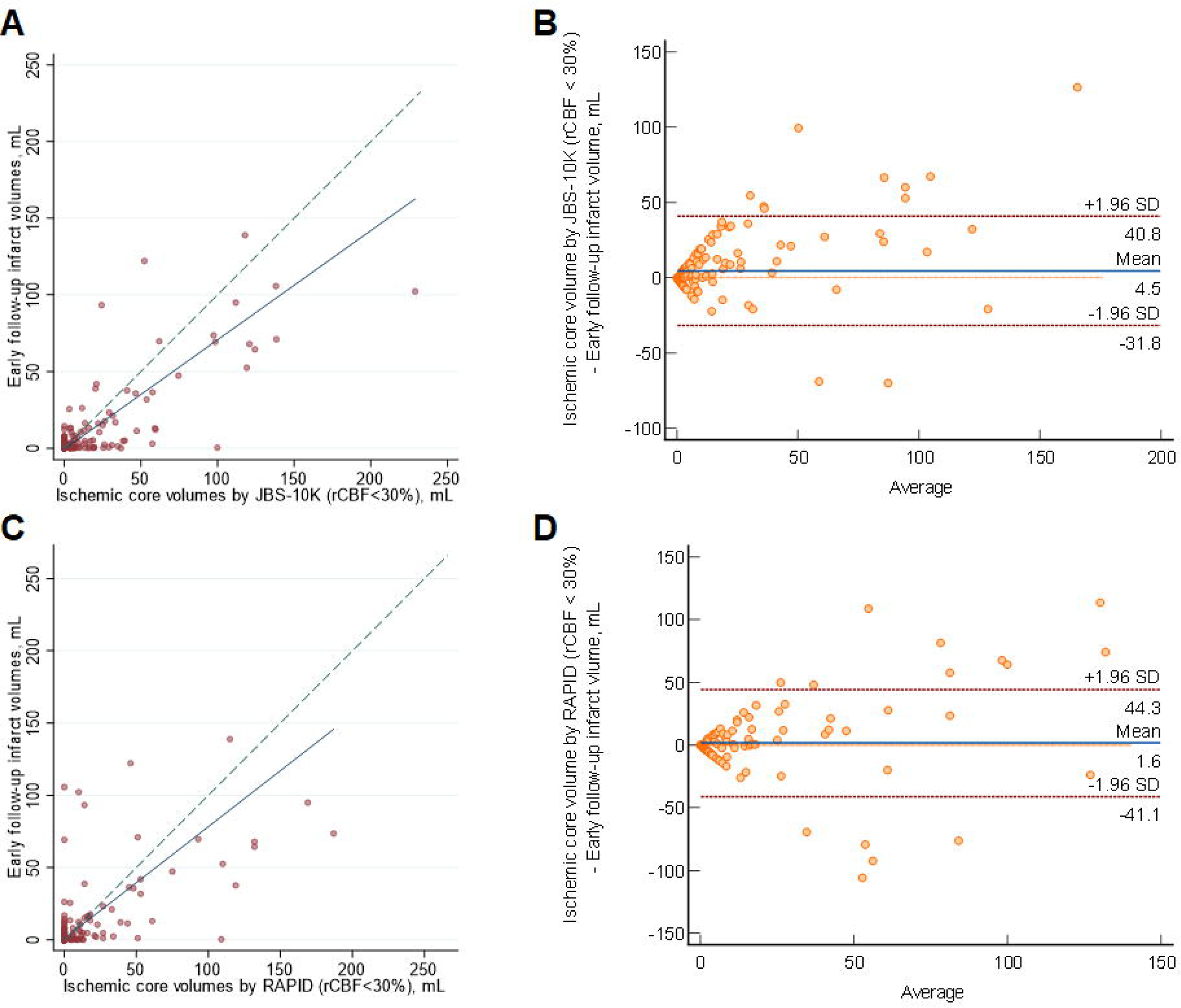
Comparison of early follow-up infarct volume on diffusion-weighted image and ischemic core volumes by JBS-10K and RAPID. (A) Scatter plot illustrating follow-up infarct volumes and ischemic core volumes as determined by JBS-10K, with a concordance correlation coefficient of 0.753, and the slope and intercept of the reduced major axis being 0.706 and −0.598, respectively. (B) Bland-Altman plot for the analysis of agreement in between follow-up infarct volumes and ischemic core volumes by JBS-01K. (C) Scatter plot illustrating follow-up infarct volumes and ischemic core volumes as determined by RAPID, with a concordance correlation coefficient of 0.736, and the slope and intercept of the reduced major axis being 0.633 and 0.804, respectively. (D) Bland-Altman plot for the analysis of agreement in between follow-up infarct volumes and ischemic core volumes by RAPID. The green dotted line represents the line of perfect concordance, while the blue line denotes the reduced major axis. For B and D, the blue solid line and the red dotted lines represent the mean difference and the limits of agreement between follow-up infarct volume and ischemic core volumes calculated by JBS-10K (B) and RAPID (D).

When dividing patients by early follow-up infarct volume (<20 mL [n=193] vs. ≥20 mL [n=24]), both packages exhibited similarly poor agreement with early follow-up infarct volume (ρ = 0.196 for JBS-10K and ρ = 0.181 for RAPID, respectively; Supplementary Figure 3). In patients whose early follow-up infarct volume was ≥20 mL, both JBS-10K (ρ = 0.497; 95% CI, 0.213 to 0.704) and RAPID demonstrated fair agreement (ρ = 0.438; 95% CI, 0.182 to 0.639). Setting the rCBF threshold to <26% improved the correlation between early follow-up infarct volume and ischemic core volumes determined by JBS-10K, as shown by both the concordance correlation coefficient and Bland-Altman analysis (Supplementary Figure 4).

### Mismatch volume analysis and application of clinical trial’s criteria

The median mismatch volumes determined by JBS-10K and RAPID were 23.26 (IQR 0 to 85.33) and 23 (0 to 101), respectively, and there was substantial agreement between the mismatch volumes determined by JBS-10K and RAPID (ρ = 0.774; 95% CI 0.735 to 0.808; Figure 4A). The mean difference in mismatch volume between the two software tools was 13.85 mL (95% CI, 8.80 to 18.90 mL; P = 0.01; Figure 4B).

**Figure 4.**
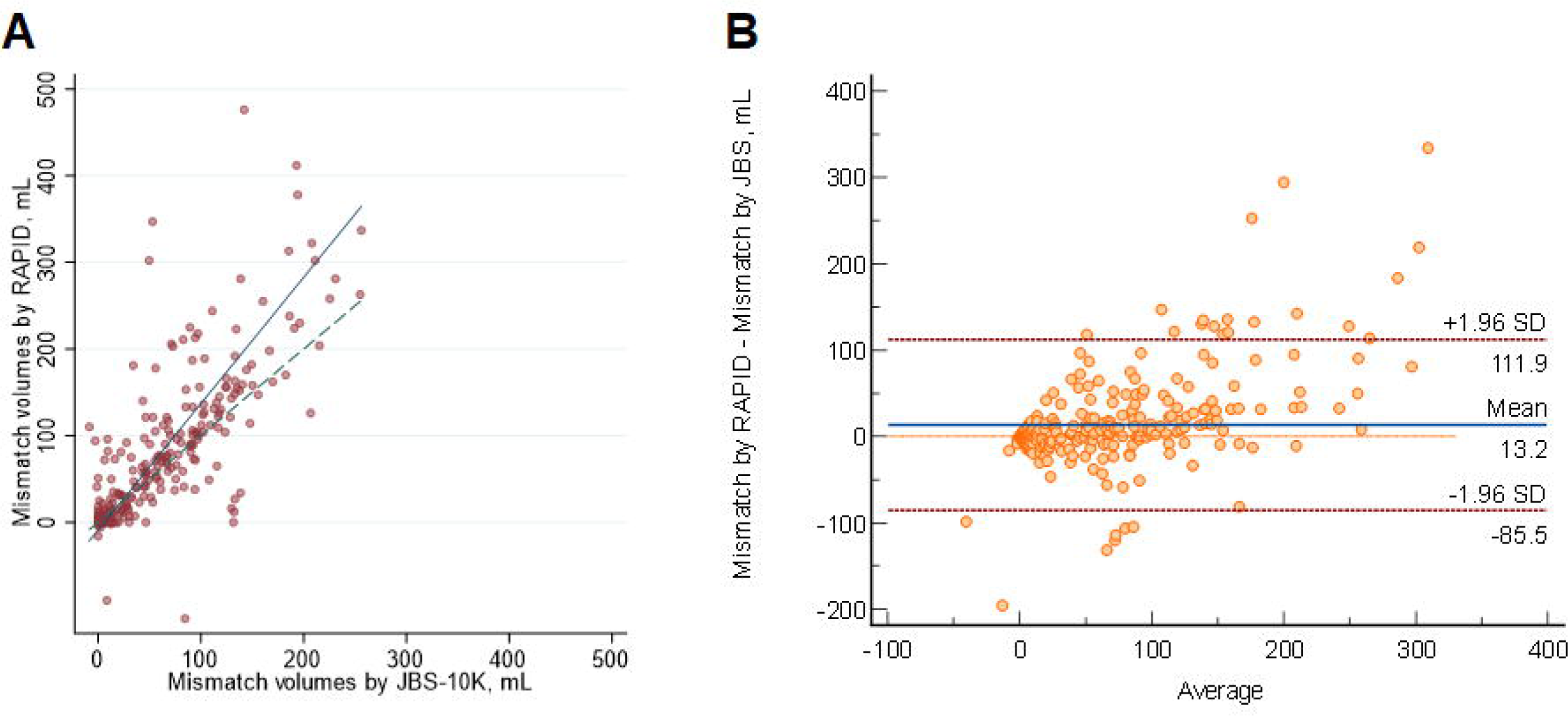
Comparison of mismatch volumes by JBS-10K and RAPID. (A) Scatter plot depicting mismatch volumes as measured by JBS-10K and RAPID, with a slope of 1.425 and an intercept of −6,783 on the reduced major axis. The green dotted line represents the line of perfect concordance, the blue line denotes the reduced major axis. (B) Bland-Altman plot illustrating the agreement in mismatch volumes between JBS-10K and RAPID. The blue line represents the mean difference, and the red dotted lines denote the limits of agreement.

When considering early infarct volume as a standard reference and categorizing ischemic core volumes by JBS-10K and RAPID according to the DAWN trial’s criteria, similar Cohen’s kappa values were observed for JBS-10K (0.55) and RAPID (0.51). However, in the medium-sized ischemic core volume groups (20–30 mL and 30–50 mL), the ischemic core volumes determined by both JBS-10K and RAPID exhibited poor agreement with early infarct volumes.

## DISCUSSION

We observed a comparable estimation of the ischemic core and hypoperfused area volume between RAPID and JBS-10K, a newly developed CTP analysis package utilizing a delay-insensitive algorithm, based on 327 ischemic stroke patients who had CTP scans within 24 hours from their last known well. Furthermore, both JBS-10K and RAPID demonstrated a comparable correlation of ischemic core volumes with early DWI infarct volumes. Additionally, we observed that the default threshold of rCBF < 30% tends to overestimate ischemic core volumes in comparison with early follow-up infarct volumes on DWI.

We observed an excellent agreement between ischemic core volumes calculated by RAPID and JBS-10K. This correlation remained high even after excluding patients with ischemic core volumes of zero as determined by either JBS-10K or RAPID. Additionally, hypoperfused tissue volumes identified using the default settings of both software tools also demonstrated substantial agreement. Consequently, mismatch volumes calculated by both software tools exhibited a substantial agreement. When the image criteria from the DAWN trial was applied,^2^ ischemic core volumes calculated by the two packages showed overall good agreement with early follow-up infarct volumes. However, for medium-sized ischemic core volumes (20-30 mL and 30-50 mL), the estimated parameters differed substantially compared with early infarct volumes on DWI. These findings suggest that ischemic core volume in CTP scans, analyzed by automated software packages, should be interpreted with caution in patients with medium-sized ischemic core volumes.

DWI is the method of choice when it comes to assessing ischemic core.^21^ Our results showed that automated CTP analysis packages have a tendency to overestimate ischemic core volume, consistent with previous studies.^22–24^ Compared to JBS-10K, RAPID was more likely to estimate the ischemic core as zero even when the ischemic core exceeded 20 mL. The discrepancy may result from differences in preprocessing images to correct patient motion and post-processing to reduce noise in both rCBF and Tmax maps. Moreover, our results contrast with a prior study demonstrating that CTP analysis packages tend to underestimate follow-up infarct volume on various imaging modalities such as DWI, fluid-attenuated inversion recovery image, and non-contrast CT.^25^ This discrepancy may result from different study populations: consecutive patients presenting at the hospital within 24 hours of onset versus patients undergoing mechanical thrombectomy. In addition, shorter time intervals from the last known well to CTP scans (median 168 vs. 402 min) and from CTP to follow-up images (median 0.7 vs. 18.7 hours) in our study may have led to smaller follow-up infarct volumes on DWI, subsequently leading to the overestimation of ischemic core volume in CTP scans by software tools. Given that DWI is the gold standard for measuring ischemic core and CTP scans are screening tools to select patients who may benefit from intervention, a more stringent rCBF threshold may be required to accurately estimate infarct core, as shown in our analysis employing an rCBF threshold of 26%, which in turn ultimately provides opportunities for more patients with ischemic stroke to regain functional independence.

The interpretation of our findings must be contextualized within the inherent limitations of this research. Conducted as a retrospective analysis at a single stroke center, our study’s observational nature is susceptible to the influence of unmeasured confounding variables. The software utilized for analysis reflects the versions available at the time of the study, acknowledging that subsequent updates and modifications could affect the applicability of our results. Moreover, the management of patients included in this study was guided by the outcomes from the RAPID software, introducing a potential source of bias. Furthermore, we utilized follow-up infarct volumes that were automatically segmented by validated software. This approach may limit comparing our results with prior studies that utilized a threshold for apparent diffusion coefficients of 620 ×10^−6^ mm^2^/s.

## CONCLUSIONS

In conclusion, the perfusion parameters from the JBS-10K and RAPID were documented to be substantially consistent. In addition, ischemic core volumes calculated by JBS-10K and RAPID at the default rCBF threshold of <30% also showed excellent agreement with early follow-up infarct volumes on DWI. However, CTP parameters should be interpreted with caution when making recanalization treatment decisions.

## Contributors

N-HK, SYH, W-SR and BJK made substantial contributions to the conception and design of the work. Data acquisition was performed by LS, M-SK, SHB, CJ, and BJK. N-HK, GP, and J-HP performed the data analysis. Interpretation of the data was carried out by DK, W-SR, and BJK. N-HK, SYH, W-SR and BJK drafted the manuscript, and all of the other authors revised it critically for important intellectual content. All authors approved the final version to be published. They agree to be accountable for all aspects of the work in ensuring that questions related to the accuracy or integrity of any part of the manuscript are appropriately investigated and resolved. Guarantor: W-SR and BJK.

## Patient consent for publication

Not applicable.

## Ethics approval

The study protocol was approved by the institutional review board of Seoul National University Bundang hospital [IRB# B-1710-429-102], and patients or their legal representatives provided a written informed consent.

## Data availability statement

Data are available upon reasonable request.

## Data Availability

All data produced in the present study are available upon reasonable request to the authors.

---

**Table 2.** Comparison of ischemic core volumes by JBS-10K and RAPID versus early follow-up infarct volume stratifying by the DAWN trial (n = 218)

|  |  | JBS-10K |  |  |  |
| --- | --- | --- | --- | --- | --- |
|  |  | 0-20 mL | 20-30 mL | 30-50 mL | > 50 mL |
| Follow-up infarct volume on DWI | 0-20 mL | 175 | 7 | 7 | 4 |
|  | 20-30 mL | 2 | 1 | 1 | 0 |
|  | 30-50 mL | 0 | 2 | 2 | 3 |
|  | > 50 mL | 0 | 1 | 0 | 13 |
|  |  | RAPID |  |  |  |
| Follow-up infarct volume on DWI | 0-20 mL | 182 | 5 | 3 | 3 |
|  | 20-30 mL | 2 | 1 | 1 | 0 |
|  | 30-50 mL | 1 | 0 | 2 | 4 |
|  | > 50 mL | 4 | 0 | 1 | 9 |

DAWN=DWI or CTP Assessment with Clinical Mismatch in the Triage of Wake-Up and Late Presenting Strokes Undergoing Neurointervention with Trevo; DWI=diffusion-weighted image.

## References

1. Abdalkader M, Siegler JE, Lee JS, Yaghi S, Qiu Z, Huo X, Miao Z, Campbell BCV, Nguyen TN. Neuroimaging of Acute Ischemic Stroke: Multimodal Imaging Approach for Acute Endovascular Therapy. J Stroke. 2023;25:55–71.

2. Jovin TG, Nogueira RG, Investigators D. Thrombectomy 6 to 24 Hours after Stroke. N Engl J Med. 2018;378:1161–1162.

3. Albers GW, Marks MP, Kemp S, Christensen S, Tsai JP, Ortega-Gutierrez S, McTaggart RA, Torbey MT, Kim-Tenser M, Leslie-Mazwi T, et al. Thrombectomy for Stroke at 6 to 16 Hours with Selection by Perfusion Imaging. N Engl J Med. 2018;378:708–718.

4. Kim BJ, Menon BK, Kim JY, Shin DW, Baik SH, Jung C, Han MK, Demchuk A, Bae HJ. Endovascular Treatment After Stroke Due to Large Vessel Occlusion for Patients Presenting Very Late From Time Last Known Well. JAMA Neurol. 2020;78:21–29.

5. Sarraj A, Grotta JC, Pujara DK, Shaker F, Tsivgoulis G. Triage imaging and outcome measures for large core stroke thrombectomy - a systematic review and meta-analysis. J Neurointerv Surg. 2020;12:1172–1179.

6. Lim NE, Chia B, Bulsara MK, Parsons M, Hankey GJ, Bivard A. Automated CT Perfusion Detection of the Acute Infarct Core in Ischemic Stroke: A Systematic Review and Meta-Analysis. Cerebrovasc Dis. 2023;52:97–109.

7. Yedavalli V, Kihira S, Shahrouki P, Hamam O, Tavakkol E, McArthur M, Qiao J, Johanna F, Doshi A, Vagal A, et al. CTP-based estimated ischemic core: A comparative multicenter study between Olea and RAPID software. J Stroke Cerebrovasc Dis. 2023;32:107297.

8. Ballout AA, Oh SY, Huang B, Patsalides A, Libman RB. Ghost infarct core: A systematic review of the frequency, magnitude, and variables of CT perfusion overestimation. J Neuroimaging. 2023;33:716–724.

9. Fainardi E, Busto G, Rosi A, Scola E, Casetta I, Bernardoni A, Saletti A, Arba F, Nencini P, Limbucci N, et al. T(max) Volumes Predict Final Infarct Size and Functional Outcome in Ischemic Stroke Patients Receiving Endovascular Treatment. Ann Neurol. 2022;91:878–888.

10. Koopman MS, Berkhemer OA, Geuskens R, Emmer BJ, van Walderveen MAA, Jenniskens SFM, van Zwam WH, van Oostenbrugge RJ, van der Lugt A, Dippel DWJ, et al. Comparison of three commonly used CT perfusion software packages in patients with acute ischemic stroke. J Neurointerv Surg. 2019;11:1249–1256.

11. Venketasubramanian N, Yoon BW, Pandian J, Navarro JC. Stroke Epidemiology in South, East, and South-East Asia: A Review. J Stroke. 2017;19:286–294.

12. Kim BJ, Lee KM, Lee SH, Kim HG, Kim EJ, Heo SH, Chang DI, Kim JS. Ethnic Differences in Intracranial Artery Tortuosity: A Possible Reason for Different Locations of Cerebral Atherosclerosis. J Stroke. 2018;20:140–141.

13. Kang DW, Kim DY, Kim J, Baik SH, Jung C, Singh N, Song JW, Bae HJ, Kim BJ. Emerging Concept of Intracranial Arterial Diseases: The Role of High Resolution Vessel Wall MRI. J Stroke. 2024;26:26–40.

14. Qiao Y, Suri FK, Zhang Y, Liu L, Gottesman R, Alonso A, Guallar E, Wasserman BA. Racial Differences in Prevalence and Risk for Intracranial Atherosclerosis in a US Community-Based Population. JAMA Cardiol. 2017;2:1341–1348.

15. Sacco RL, Kargman DE, Gu Q, Zamanillo MC. Race-ethnicity and determinants of intracranial atherosclerotic cerebral infarction. The Northern Manhattan Stroke Study. Stroke. 1995;26:14–20.

16. Kim BJ, Park JM, Kang K, Lee SJ, Ko Y, Kim JG, Cha JK, Kim DH, Nah HW, Han MK, et al. Case characteristics, hyperacute treatment, and outcome information from the clinical research center for stroke-fifth division registry in South Korea. J Stroke. 2015;17:38–53.

17. Ko Y, Lee S, Chung JW, Han MK, Park JM, Kang K, Park TH, Park SS, Cho YJ, Hong KS, et al. MRI-based Algorithm for Acute Ischemic Stroke Subtype Classification. J Stroke. 2014;16:161–172.

18. Kudo K, Sasaki M, Yamada K, Momoshima S, Utsunomiya H, Shirato H, Ogasawara K. Differences in CT perfusion maps generated by different commercial software: quantitative analysis by using identical source data of acute stroke patients. Radiology. 2010;254:200–209.

19. Ryu WS, Kang YR, Noh YG, Park JH, Kim D, Kim BC, Park MS, Kim BJ, Kim JT. Acute Infarct Segmentation on Diffusion-Weighted Imaging Using Deep Learning Algorithm and RAPID MRI. J Stroke. 2023;25:425–429.

20. Lin LI. A concordance correlation coefficient to evaluate reproducibility. Biometrics. 1989;45:255–268.

21. van Everdingen KJ, van der Grond J, Kappelle LJ, Ramos LM, Mali WP. Diffusion-weighted magnetic resonance imaging in acute stroke. Stroke. 1998;29:1783–1790.

22. Garcia-Tornel A, Campos D, Rubiera M, Boned S, Olive-Gadea M, Requena M, Ciolli L, Muchada M, Pagola J, Rodriguez-Luna D, et al. Ischemic Core Overestimation on Computed Tomography Perfusion. Stroke. 2021;52:1751–1760.

23. Sarraj A, Campbell BCV, Christensen S, Sitton CW, Khanpara S, Riascos RF, Pujara D, Shaker F, Sharma G, Lansberg MG, et al. Accuracy of CT Perfusion-Based Core Estimation of Follow-up Infarction: Effects of Time Since Last Known Well. Neurology. 2022;98:e2084–e2096.

24. Boned S, Padroni M, Rubiera M, Tomasello A, Coscojuela P, Romero N, Muchada M, Rodriguez-Luna D, Flores A, Rodriguez N, et al. Admission CT perfusion may overestimate initial infarct core: the ghost infarct core concept. J Neurointerv Surg. 2017;9:66–69.

25. Pisani L, Haussen DC, Mohammaden M, Perry da Camara C, Jillella DV, Rodrigues GM, Bouslama M, Al-Bayati A, Prater A, Liberato B, et al. Comparison of CT Perfusion Software Packages for Thrombectomy Eligibility. Ann Neurol. 2023;94:848–855.

