## Supplementary Material for "Comparison of two automated CT perfusion software packages in patients with ischemic stroke presenting within 24 hours of onset"

**Supplementary Figure 1. Comparison of ischemic core volumes by JBS-10K and RAPID in patients whose ischemic core volume was not zero by both JBS-10K and RAPID**

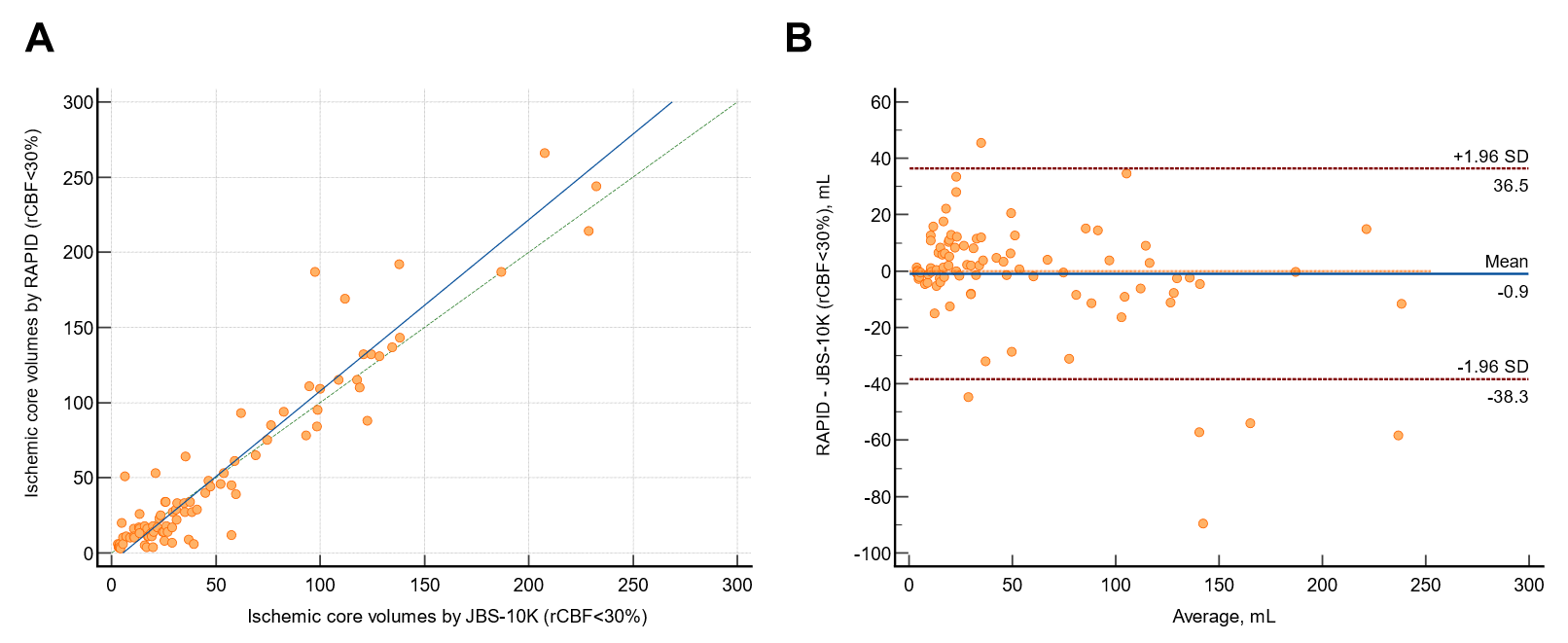

(A) Scatter plot depicting ischemic core volumes as measured by JBS-10K and RAPID, with a slope of 1.140 and an intercept of -6.257 on the reduced major axis. The green dotted line represents the line of perfect concordance, the blue line denotes the reduced major axis. (B) Bland-Altman plot illustrating the agreement in ischemic core volumes between JBS-10K and RAPID. The blue line represents the mean difference, and the red dotted lines denote the limits of agreement.

**Supplementary Figure 2. Comparison of ischemic core volumes and hypoperfused tissue volumes by JBS-10K and RAPID in patients receiving endovascular treatment (n = 122)**

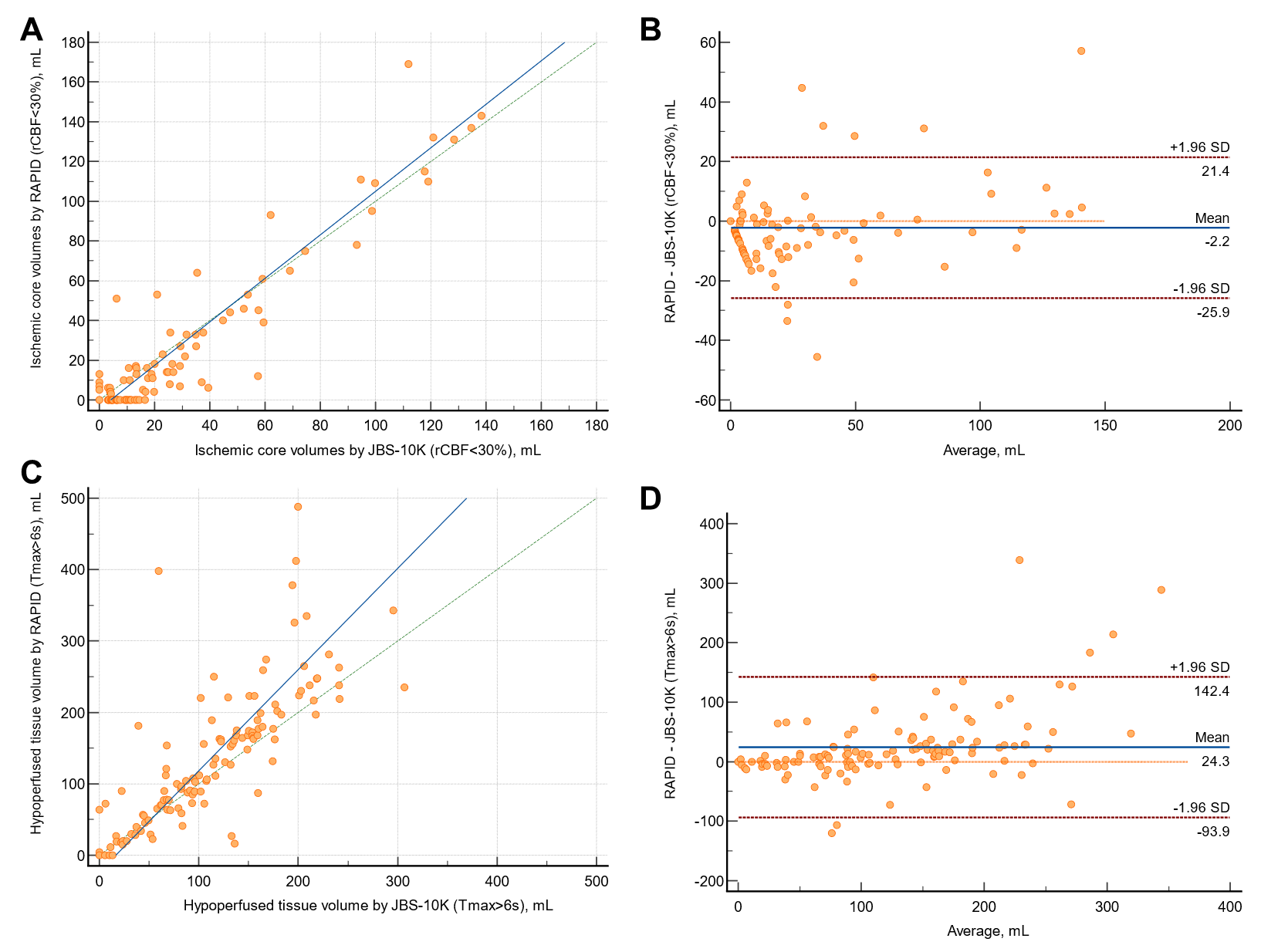

(A) Scatter plot illustrating ischemic core volumes as determined by JBS-10K and RAPID, with a concordance correlation coefficient of 0.940, and the slope and intercept of the reduced major axis being 1.095 and -4.511, respectively. (B) Bland-Altman plot for analyzing agreement in ischemic core volumes. (C) Scatter plot illustrating hypoperfused tissue volumes as determined by JBS-10K and RAPID, with a concordance correlation coefficient of 0.709, and the slope and intercept of the reduced major axis being 1.417 and -23.422, respectively. (D) Bland-Altman plot for analyzing agreement in hypoperfused tissue volumes. The green dotted line represents the line of perfect concordance, the blue line denotes the reduced major axis. For the B and D, the blue solid line alongside the red dotted lines indicate the mean difference and limits of agreement between JBS-10K and RAPID, respectively.

**Supplementary Figure 3. Comparison of early follow-up infarct volumes and ischemic core volumes by JBS-10K and RAPID after stratified by follow-up infarct volume (< 20 mL versus ≥ 20 mL)**

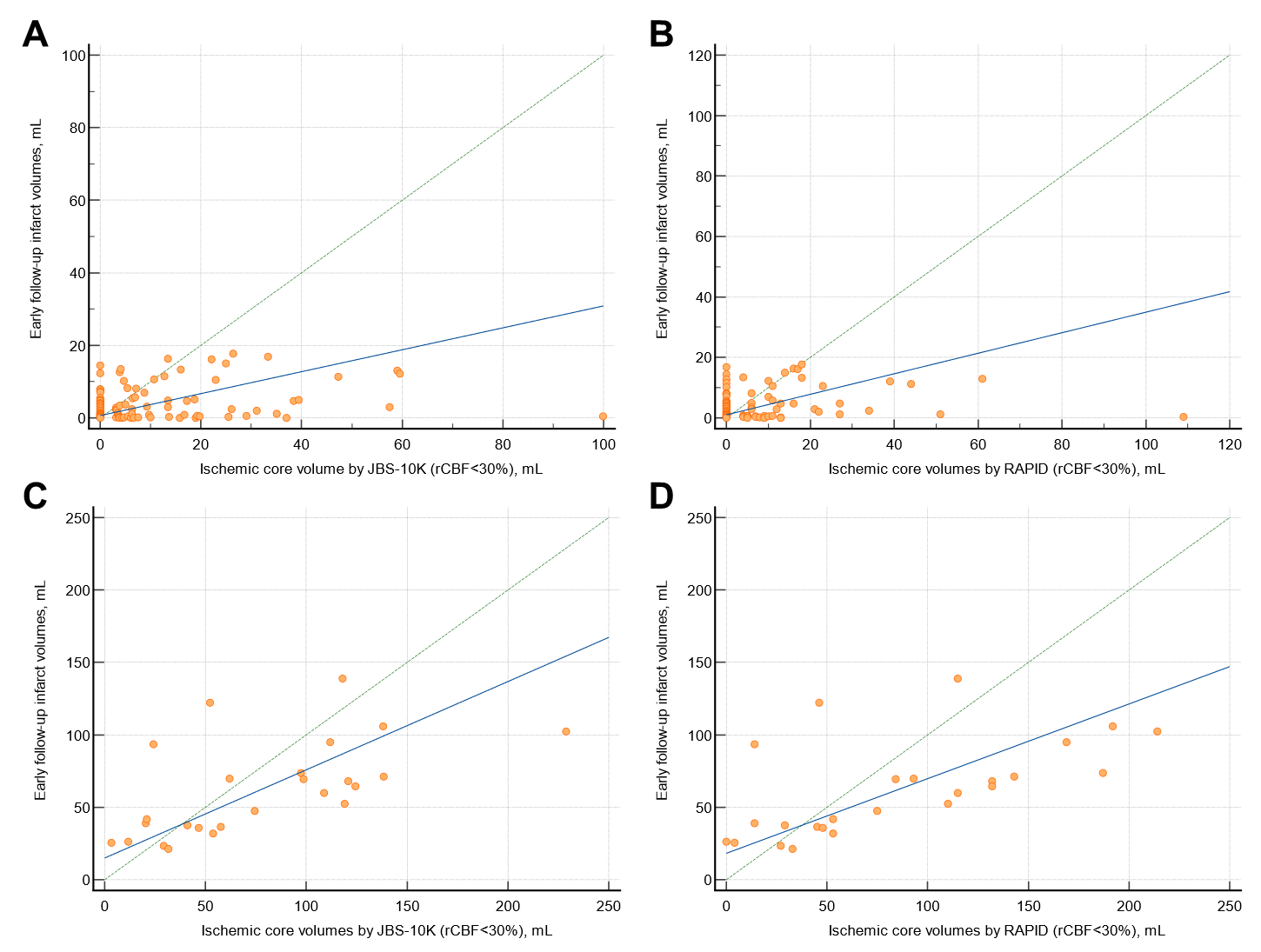

In patients with small infarct volumes on follow-up diffusion weighted image (< 20 mL) (A) Scatter plots showing early follow infarct volumes and ischemic core volumes by JBS-10K, with concordance correlation coefficient of 0.195, and the slope and intercept of the reduced major axis being 0.302 and 0.650, respectively. (B) Scatter plots showing early follow infarct volumes and ischemic core volumes by RAPID, with concordance correlation coefficient of 0.181, and the slope and intercept of the reduced major axis being 0.339 and 1.017, respectively. In patients with large infarct volumes on follow-up diffusion weighted image (≥20 mL) (C) Scatter plots showing early follow infarct volumes and ischemic core volumes by JBS-10K, with concordance correlation coefficient of 0.497, and the slope and intercept of the reduced major axis being 0.609 and 14.955, respectively. (D) Scatter plots showing early follow infarct volumes and ischemic core volumes by RAPID, with concordance correlation coefficient of 0.438, and the slope and intercept of the reduced major axis being 0.516 and 18.194, respectively.

**Supplementary Figure 4. Comparison of early follow-up infarct volumes and ischemic core volumes by JBS-10K at the threshold of relative cerebral blood flow (rCBF) of 26%**

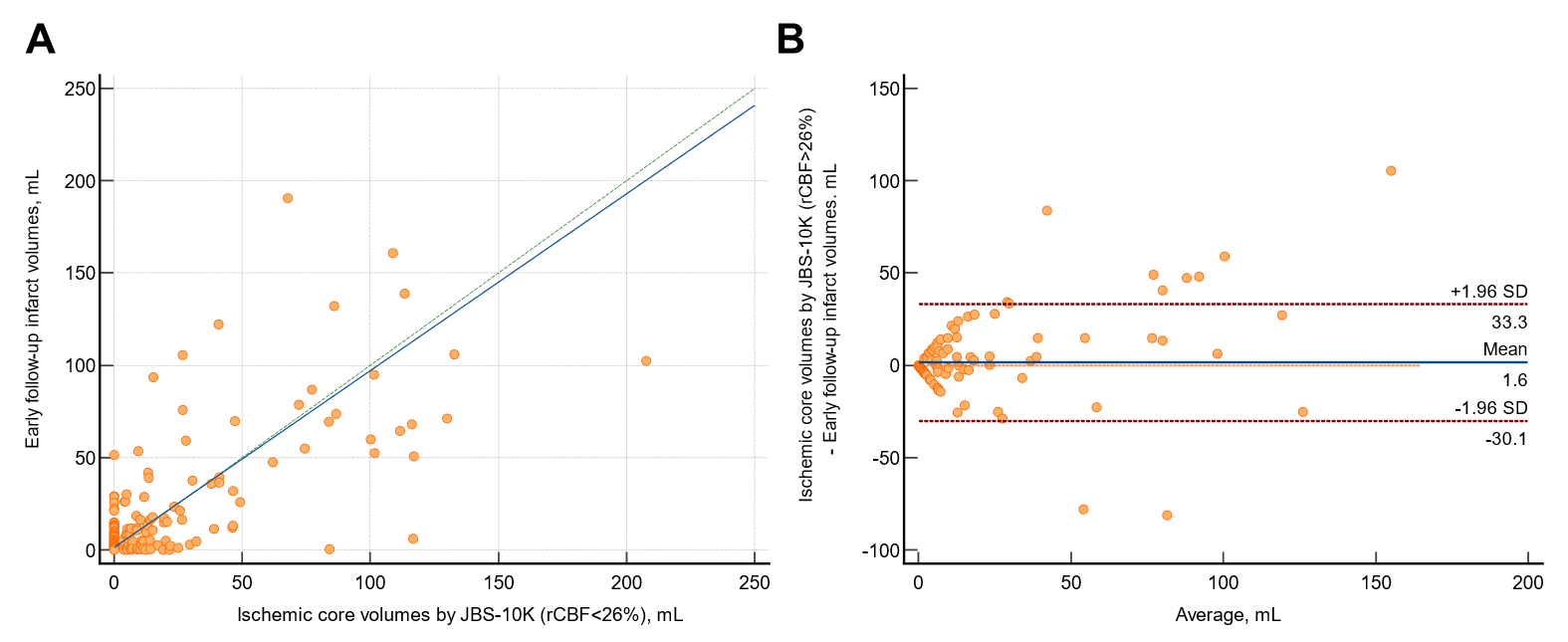

(A) Scatter plot depicting ischemic core volumes as measured by JBS-10K at the threshold of rCBF <2 6%, with concordance correlation coefficient of 0.795, and the slope and intercept of the reduced major axis being 0.798 and 0.706, respectively. The green dotted line represents the line of perfect concordance, the blue line denotes the reduced major axis. (B) Bland-Altman plot illustrating the agreement between follow-up infarct volumes and ischemic core volumes by JBS-10K at the threshold of rCBF < 26%. The blue line represents the mean difference, and the red dotted lines denote the limits of agreement.
